# Portable Breathing Monitoring with Phase-Resolved Airflow Dynamics Enabled by a Dual-Response Flexible PZT Sensor

**DOI:** 10.64898/2026.02.09.26345795

**Authors:** Minyu Li, Jun Aoyama, Yuchao Wu, Tomomi Uchiyama, Kosho Yoshikawa, Toshiki Mano, Yuxi Song, Hedong Zhang

## Abstract

Respiratory monitoring in daily-life settings is important for health assessment, yet extracting physiologically interpretable information from breathing signals under natural conditions remains challenging, as breathing is inherently dynamic and strongly modulated by behavior. Here, a portable breathing monitoring device based on a flexible lead zirconate titanate sensor is developed to address this challenge. By exploiting polarity-opposed piezoelectric and pyroelectric responses through sensor orientation, the recorded breathing waveform exhibits a characteristic dual-component structure, consisting of a narrow transient spike followed by a broad quasi-steady peak within each breathing phase. This intrinsic waveform structure enables phase-resolved quantification of how breathing effort is distributed between transient and quasi-steady components during inhalation and exhalation. Pilot measurements in healthy subjects and patients with chronic obstructive pulmonary disease or asthma reveal systematic shifts toward transient-enhanced breathing in patients, providing clearer differentiation than conventional descriptors based on breathing duration or amplitude. By transforming complex breathing dynamics into stable and physiologically meaningful signal components under daily-life conditions, this dual-response sensing approach enables more robust access to function-related changes in natural breathing.

## 1. Introduction

Respiration is one of the most fundamental vital signs and forms a tightly coupled regulatory loop with autonomic and cardiovascular systems.^[1–4]^ In addition to respiratory diseases such as chronic obstructive pulmonary disease (COPD) and asthma,^[5,6]^ respiratory rate and pattern changes have been shown to be closely associated with cardiovascular function and overall systemic health.^[1–3,7]^ Previous studies have indicated that subtle alterations in respiratory rate or detailed breathing patterns may precede detectable changes in conventional clinical indicators such as heart rate and blood pressure, suggesting respiration as a sensitive early marker of health deterioration.^[2,8–10]^ Therefore, respiratory monitoring, particularly that can be performed repeatedly and naturally in daily life, is of great importance for long-term health assessment and more personalized clinical management.^[1,11,12]^

At present, spirometry remains the clinical gold standard for evaluating respiratory function. By directly measuring airflow under controlled conditions, it provides quantitative indices such as forced expiratory volume and vital capacity that are well established in clinical practice.^[5,6,13]^ However, spirometry relies on forced breathing tasks, strict subject cooperation, and bulky instrumentation, which fundamentally limits its applicability for repeated measurements and assessment under natural daily-life conditions.^[1,11,12,14]^ As a result, despite the recognized clinical importance of respiration, detailed respiratory information is rarely captured outside clinical or laboratory settings.

To overcome these limitations, a variety of approaches for respiratory monitoring in daily-life settings have been actively developed in recent years.^[1,11,12,15,16]^ These approaches can be broadly classified into indirect, non–airflow-based methods and direct airflow-sensing methods. Non–airflow-based approaches infer respiration from body motion, most commonly chest or abdominal movement, and are typically implemented using inertial measurement units or strain-based wearable sensors.^[17–23]^ These methods enable continuous and unobtrusive monitoring and have therefore been widely explored for long-term use. However, because the primary measured signals are mechanical body movements rather than respiratory airflow, non–airflow-based methods are intrinsically less sensitive to airflow-related physiological properties such as airway resistance, lung compliance, and flow limitation, which are central to respiratory pathophysiology.^[1,5,6]^

Direct airflow sensing, in principle, offers higher physiological relevance and closer correspondence to spirometric measurements.^[1,15,24]^ Accordingly, a variety of portable and wearable airflow sensors have been proposed in recent years.^[25–29]^ To date, most airflow-based respiratory monitoring studies have focused on steady-state or quasi-steady parameters, such as respiratory rate, tidal volume, or peak expiratory flow, which primarily characterize the overall breathing rate and depth.^[1,11,30,31]^ However, airflow during everyday natural (tidal) breathing is strongly modulated by behavior and inherently time-varying rather than steady. In particular, transitions between inhalation and exhalation involve rapid airflow changes governed by airway mechanics, lung compliance, and neuromuscular control.^[5–7,32–36]^ Emerging evidence indicates that dynamic features associated with inhalation–exhalation phase transitions may offer increased sensitivity to respiratory instability and early deterioration than classical steady-state metrics such as respiratory rate.^[37]^ Despite their physiological relevance, such transient airflow features have received relatively little attention in daily-life breathing monitoring. This is partly because conventional sensing approaches are not specifically optimized to reliably capture fast, transient airflow-related phenomena under unconstrained conditions.

From a physical perspective, breathing airflow inherently involves coupled mechanical and thermal effects, arising from pressure fluctuations and the temperature difference between inhaled and exhaled air. Capturing both aspects, rather than either one alone, is therefore expected to yield richer waveform features that reflect not only steady-state but also dynamic characteristics of airflow. Motivated by this consideration, we propose a portable breathing monitoring device based on a flexible lead zirconate titanate (PZT) sensor that simultaneously exploits piezoelectric and pyroelectric responses. Rather than allowing the two effects to merge into an indistinguishable superposition, their polarities are intentionally arranged to preserve mutually discernible contributions within a single waveform. As a result, the device produces waveforms that differ qualitatively from those of conventional airflow-sensing devices, enabling the initial transient and subsequent quasi-steady phases of each inhalation and exhalation cycle to be distinctly represented under natural oral breathing conditions. By transforming complex, time-varying airflow dynamics into stable and separable signal components, this device enables robust, phase-resolved analysis of intra-cycle breathing behavior under daily-life conditions.

## 2. Results and Discussion

### 2.1. Sensor Selection and Structural Design

A flexible PZT sensor (**Figure 1**a), which exhibits both piezoelectric and pyroelectric responses and was used in our previous study for arterial pulse waveform measurement,^[38]^ was employed in this study. In contrast to rigid bulk PZT sensors that primarily operate in the *e*_33_ mode, the flexible sensor undergoes pronounced in-plane deformation (*e*_31_ mode) even under small pressure fluctuations,^[39]^ enabling sensitive detection of weak airflow-induced mechanical stimuli during natural breathing. Additionally, compared with widely used organic polyvinylidene fluoride (PVDF) flexible sensors, our flexible PZT sensors have been confirmed to exhibit one-order-of-magnitude higher piezoelectric sensitivity^[38]^ and comparable pyroelectric sensitivity (Note S1, Supporting Information).

**Figure 1.**
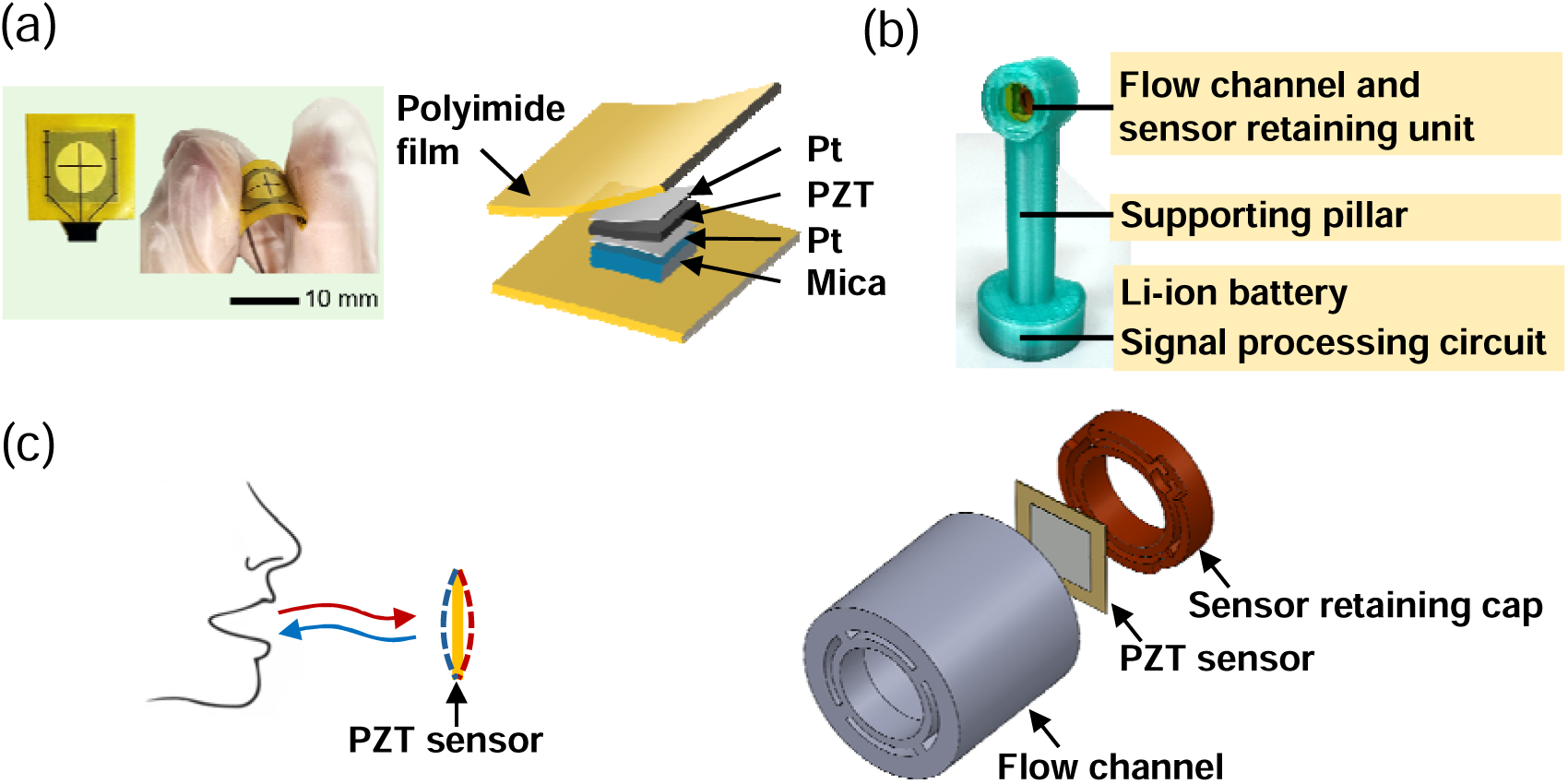
Overview of the proposed portable breathing monitoring device using a flexible PZT sensor. (a) Photographs of the flexible PZT sensor (left) and schematic illustration of its multilayer configuration (right) adapted from Ref. [38] under the terms of the Creative Commons Attribution (CC BY 4.0) license. (b) Photograph of the proposed device. (c) Schematic illustration of the sensor orientation relative to the oral breathing airflow (left) and exploded view of the sensor retaining unit (right).

Figure 1b shows a photograph of the portable device developed for monitoring natural oral breathing in this study. The device consists of a flow channel, a sensor retaining unit, a supporting pillar, and a base housing a Li-ion battery and signal processing circuit. As demonstrated in Movie S1 (Supporting Information), an attachable, commercially available mouthpiece for nebulizers is connected to the upstream side of the flow channel to guide airflow toward the sensor during natural oral breathing. The electrical charges generated by the sensor are subsequently processed and transmitted wirelessly via Bluetooth by the circuit in the base to a mobile terminal, such as a tablet or smartphone, for real-time monitoring and recording.

Among these components, the structural design of the sensing unit plays a central role in determining the signal characteristics. To ensure effective mechanical and thermal interaction with the breathing airflow, the sensor was oriented with its surface perpendicular to the airflow direction (Figure 1c, left). Because the sensor has an asymmetric multilayer configuration with a PZT-on-mica core (Figure 1a), reversing the sensor orientation changes which side of the core faces the upstream airflow. The orientation was therefore carefully designed so that the piezoelectric and pyroelectric signal components exhibit opposite polarities, preventing signal overlap and enhancing distinguishable features in the measured waveforms. The piezoelectric response is governed by the strain polarity of the PZT layer, with compressive and tensile strains generating signals of opposite polarity. As the mechanical neutral plane of the sensor lies within the mica substrate layer (Note S2, Supporting Information), the strain polarity of the PZT layer varies with the sensor orientation relative to the airflow. Specifically, when the PZT layer faces upstream, exhalation generates compressive strain and inhalation generates tensile strain, whereas reversing the sensor orientation inverts the strain polarity and consequently the polarity of piezoelectric signals. In contrast, the pyroelectric response originates from temperature variations during breathing. Exhalation-induced heating and inhalation-induced cooling produce signals of opposite polarity; however, this polarity behavior is independent of the sensor orientation. Based on these considerations and the signal polarities independently identified using purely mechanical or thermal stimuli (**Figure 2**), the sensor was oriented with the PZT layer facing upstream.

**Figure 2.**
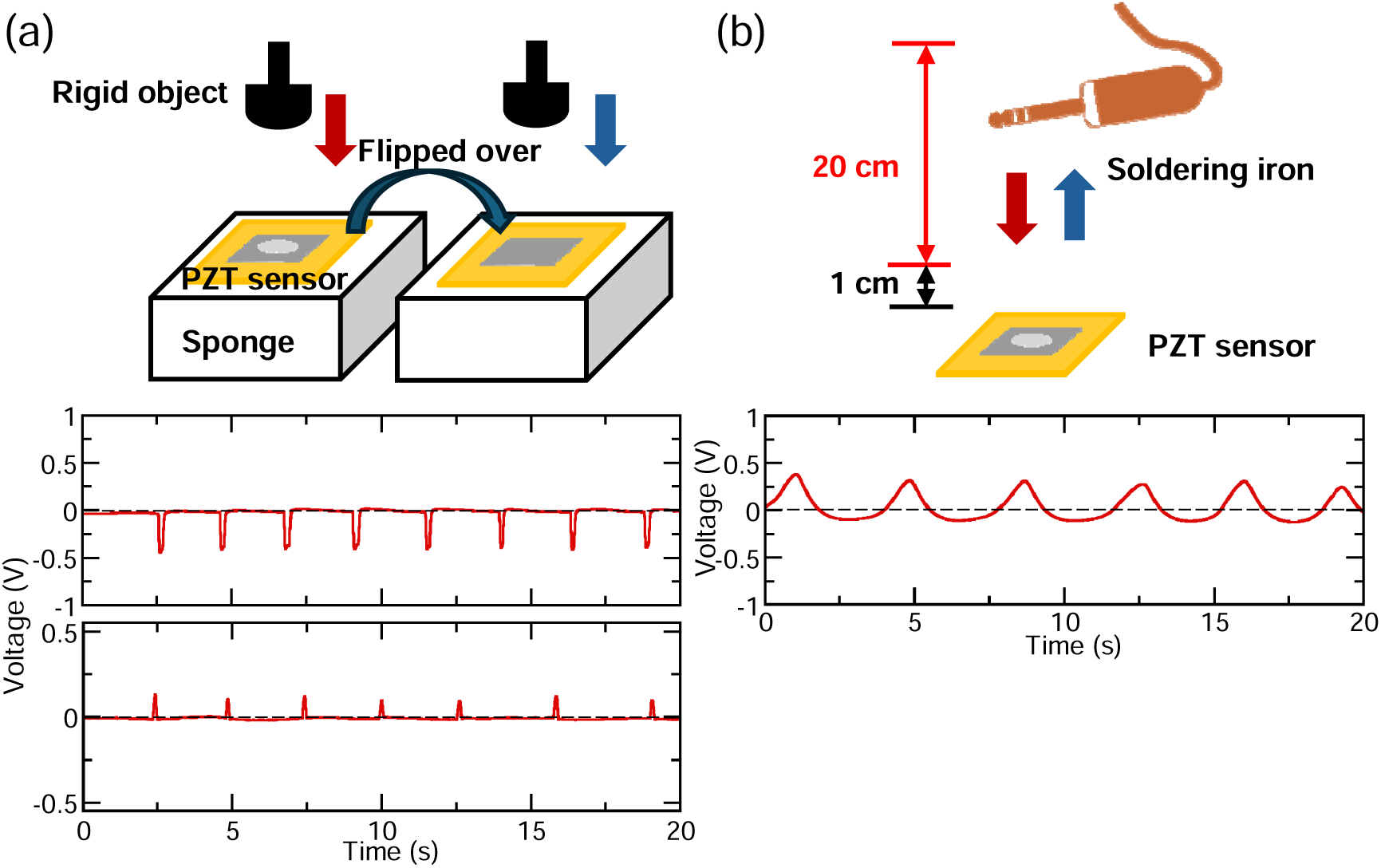
Schematic illustrations of polarity identification experiments under mechanical (a) and thermal (b) stimuli, together with the corresponding voltage outputs. Mechanical stimulus (a): the PZT sensor was placed on a soft sponge and repeatedly pressed using a handheld rounded rigid object, with the sensor tested in both original and inverted orientations to confirm signal polarity reversal. Thermal stimulus (b): a soldering iron was periodically moved vertically above the sensor within a distance range of 1–21 cm over 2 s in each direction, inducing a cyclic temperature variation of the sensor surface between 25.0 and 36.4 °C, as measured by an infrared thermometer.

The flexible PZT sensor was placed in a sensor seat at the outlet of the flow channel and secured by a retaining cap (Figure 1c, right), which mechanically clamps the sensor along its perimeter and maintains its planar configuration. Four fan-shaped openings were symmetrically arranged around the sensor to serve as airflow passages. Reducing the opening size would enhance both mechanical and thermal interactions between the sensor and airflow, leading to stronger sensor signals; however, excessively small openings would increase flow resistance and compromise breathing comfort. Therefore, the opening size was designed as the minimum required to maintain smooth and unobstructed natural breathing.

### 2.2. Breathing Waveform Characteristics in Healthy Subjects

#### 2.2.1. Representative Breathing Waveform

To understand the fundamental characteristics of the oral breathing signals acquired by the proposed device, measurements were conducted for 60 s on 15 healthy subjects with no history of respiratory disease and smoking in the past 10 years (12 males and 3 females, aged 20–60s). For comparison, a commercially available flow sensor for adult oral respiratory monitoring was connected downstream of our device to simultaneously record airflow rate signals (**Figure 3**a).

**Figure 3.**
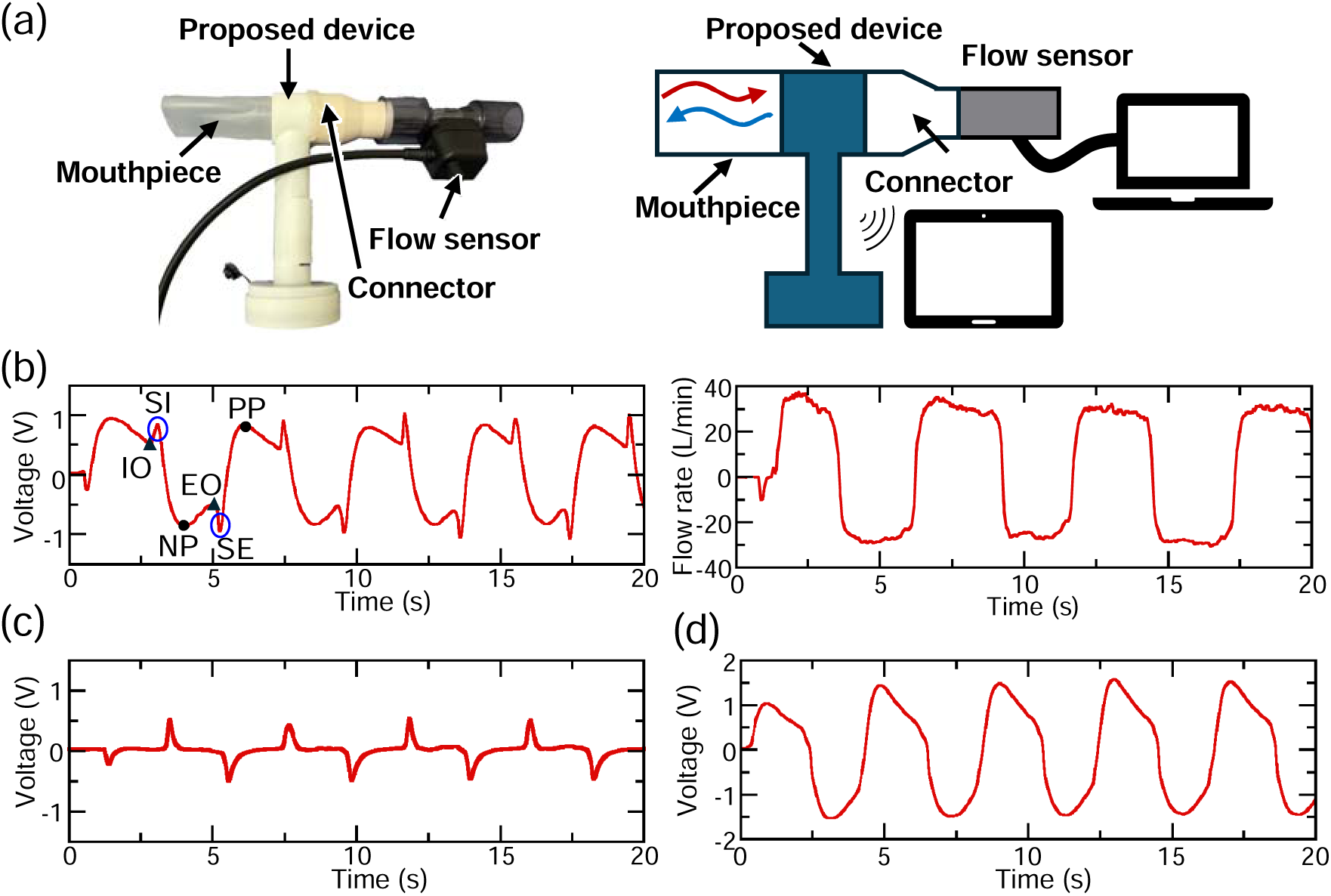
Experimental setup and representative results from healthy subjects (a,b), together with control experiment results for examining piezoelectric and pyroelectric contributions (c) and the effect of sensor orientation (d). (a) Photograph and schematic illustration of the experimental setup for oral breathing measurements. A mouthpiece is connected to the proposed device, and a commercially available flow sensor is connected downstream of the device for comparison. (b) Representative breathing waveform obtained from the proposed device (left) and the corresponding airflow rate signal recorded by the flow sensor (right) at an ambient temperature of 25.0 °C. IO: inhalation onset, SI: spike of inhalation, NP: negative peak, EO: exhalation onset, SE: spike of exhalation, PP: positive peak. (c) Breathing waveform measured at an ambient temperature of 36.3 °C. (d) Breathing waveform measured with the sensor reversed relative to the airflow direction (PZT layer facing downstream) at an ambient temperature of 25.0 °C.

Figure 3b shows a representative breathing waveform obtained from our device (left), together with the corresponding signal recorded simultaneously by the flow sensor (right). For clarity, only the initial 20 s segments of the 60 s recordings are shown to illustrate waveform features. In contrast to the flow sensor signal, the waveform acquired by our device exhibits spike-like features, highlighted by circles, together with a broader peak within each breathing phase. Each inhalation onset (IO), as recognized by the subject at the start of inhalation, was consistently observed at a local minimum in the waveform, followed sequentially by a narrow positive spike (SI) and a broad negative peak (NP). Conversely, each exhalation onset (EO) occurred at a local maximum, followed by a narrow negative spike (SE) and a broad positive peak (PP).

#### 2.2.2. Origin of Spike Features

As described above, representative breathing waveforms include one spike and one peak with opposite polarities within each inhalation or exhalation phase. Based on signal polarities independently identified using purely mechanical or thermal stimuli (Figure 2), the piezoelectric response should be positive and the pyroelectric response should be negative during inhalation, whereas the polarities should be reversed during exhalation. By directly comparing these polarity relationships with the measured breathing waveforms, the narrow spike and broad peak components can be attributed to the piezoelectric and pyroelectric responses, respectively.

To further validate this attribution, control experiments were performed to suppress the pyroelectric contribution by reducing the temperature difference between exhalation and inhalation, i.e., between body and ambient temperatures. Compared with the waveform measured at an ambient temperature of 25.0 °C (Figure 3b, left), the broad peak components nearly disappeared at an ambient temperature of 36.3 °C, close to body temperature, while the narrow spike components remained clearly observable (Figure 3c). This result directly validates the above attribution.

In addition, to confirm the influence of sensor orientation on the waveform characteristics, a comparative experiment was performed by reversing the sensor with respect to the airflow direction, i.e., with the PZT layer facing downstream. As shown in Figure 3d, the spike features observed in the normal configuration (Figure 3b, left) disappeared under the reversed orientation. This result is consistent with the expected polarity relationship between the piezoelectric and pyroelectric responses. Reversing the sensor orientation inverts the polarity of the piezoelectric response, causing it to align with that of the pyroelectric response and resulting in same-polarity signal superposition without spike features. Hence, the breathing waveforms measured by the proposed device are characterized by sensor-orientation-dependent spike features, which originate from the piezoelectric response exhibiting polarities opposite to those of the simultaneously acquired pyroelectric response.

#### 2.2.3. Relationship between Spike Features and Breathing Characteristics

The spike features observed shortly after inhalation or exhalation onset are expected to reflect rapid airflow changes during the initial transient phase of breathing. This is because the piezoelectric response, as read out by a charge amplifier, is sensitive to time-varying deformation rather than static deformation.^[40,41]^ We therefore examined the relationship between spike features and the rate of airflow change during these transient phases. Because this rate cannot be directly controlled during voluntary breathing, it was indirectly modulated in a single representative subject under three conditions: (i) natural breathing, (ii) deeper breathing at approximately the same breathing frequency, and (iii) faster breathing at approximately the same peak-to-valley amplitude. The breathing amplitude and frequency in the latter two conditions were adjusted relative to natural breathing and confirmed using simultaneously recorded flow sensor signals to remain within −6% of the intended constraint.

**Figure 4** shows the breathing waveforms measured by our device (left) and the corresponding airflow rate signals measured by the flow sensor (right). Compared with natural breathing (upper left), both deeper breathing (middle left) and faster breathing (lower left) resulted in a pronounced increase in spike amplitude. Quantitatively, the mean spike amplitude increased to approximately 2.1-fold during deeper breathing and 1.2-fold during faster breathing compared with natural breathing. Despite the different magnitudes, the observed increases in both conditions suggest that spike amplitude is more closely governed by the rate of airflow change immediately after inhalation or exhalation onset than by breathing depth or breathing rate alone.

**Figure 4.**
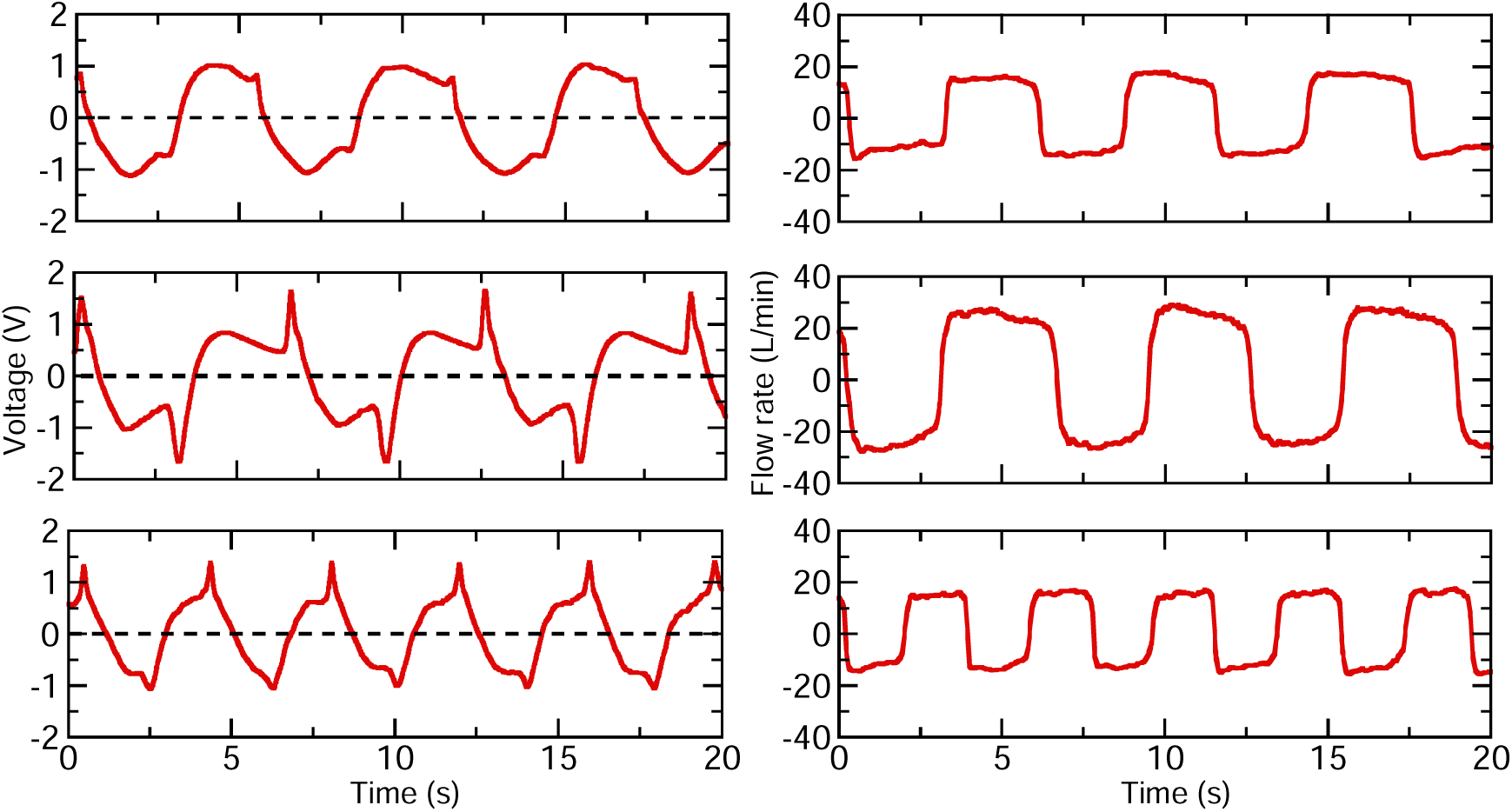
Control experiment results showing the correspondence between spike features and breathing under different depths and rates. Breathing waveforms measured by the proposed device (left) and the corresponding airflow rate signals measured by the flow sensor (right) for natural (upper), deeper (middle), and faster (lower) breathing.

### 2.3. Differences among COPD, Asthma, and Healthy Subjects

We performed a pilot comparison of breathing waveforms acquired by our device among patients with COPD (3 males and 1 female, aged 60–80s), patients with asthma (3 males and 2 females, aged 50–80s), and the previously mentioned 15 healthy subjects. Subjects were included based on availability and informed consent, without waveform-based preselection. Given the limited sample size, particularly in the patient groups, this analysis primarily focuses on characterizing natural breathing patterns in healthy individuals, using them as a reference to examine disease-related deviations rather than to establish diagnostic criteria.

#### 2.3.1. Phase-Resolved Metrics

A representative breathing waveform from patients over the initial 20 s is shown in **Figure 5** to highlight characteristic differences from healthy subjects, while waveforms from the remaining patients are provided in Figure S2 (Supporting Information). Compared with healthy subjects (Figure 3b, left), this representative patient waveform shows an enhanced spike component, with increased amplitude and temporal contribution relative to the subsequent peak component. For clarity in the following analysis, the spike component is referred to as the transient phase, and the subsequent peak component as the post-transient (quasi-steady) phase.

**Figure 5.**
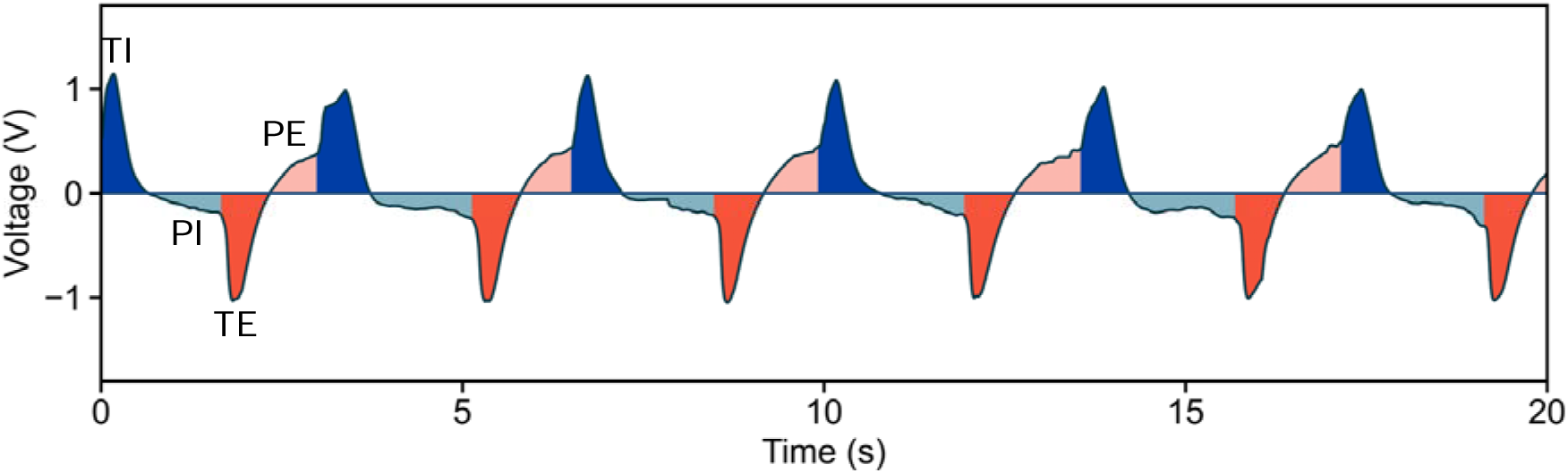
Representative breathing waveform from a COPD patient, showing characteristic differences from healthy subjects, together with the phase-resolved segmentation used in the subsequent analysis. Each breathing cycle was segmented into four phases: transient inhalation (blue), post-transient (quasi-steady) inhalation (light blue), transient exhalation (red), and post-transient (quasi-steady) exhalation (light red). These phase-resolved segments were used to compute the integral metrics *R*_TI_, *R*_PI_, *R*_TE_, and *R*_PE_.

To quantitatively characterize the relative contributions of the transient and post-transient (quasi-steady) phases, we defined phase-resolved integral ratio metrics, *R*_TI_, *R*_PI_, *R*_TE_, and *R*_PE_, as illustrated in Figure 5. Specifically, each breathing cycle was segmented into four regions corresponding to the transient and quasi-steady phases during inhalation and exhalation. The integrated area of each region was calculated and normalized by the total integrated area over the corresponding breathing cycle to obtain the metrics, which were then averaged over a 60 s recording period for each subject.

**Figure 6** shows jittered scatter plots of the phase-resolved integral ratio metrics for healthy subjects and patients with COPD or asthma. Healthy subjects exhibited a narrow and well-defined distribution across all four metrics, as highlighted by the shaded reference region. Specifically, the transient-phase ratios (*R*_TI_ and *R*_TE_) remained low, typically within 0–20%, whereas the quasi-steady-phase ratios (*R*_PI_ and *R*_PE_) accounted for a larger fraction of the breathing cycle, typically within 30–50%. In contrast, although some patients exhibited values comparable to those of healthy subjects, a subset displayed pronounced deviations beyond the healthy reference region characterized by increased transient-phase ratios (approximately 20–40%) and reduced quasi-steady-phase ratios (approximately 10–30%). Such deviations were observed slightly more frequently in the inhalation quasi-steady phase (*R*_PI_) and the exhalation transient phase (*R*_TE_) than in the inhalation transient phase (*R*_TI_) and the exhalation quasi-steady phase (*R*_PE_). This tendency was also reflected in the group-averaged values, with more pronounced differences between patients and healthy subjects for *R*_PI_ and *R*_TE_ than for *R*_TI_ and *R*_PE_.

**Figure 6.**
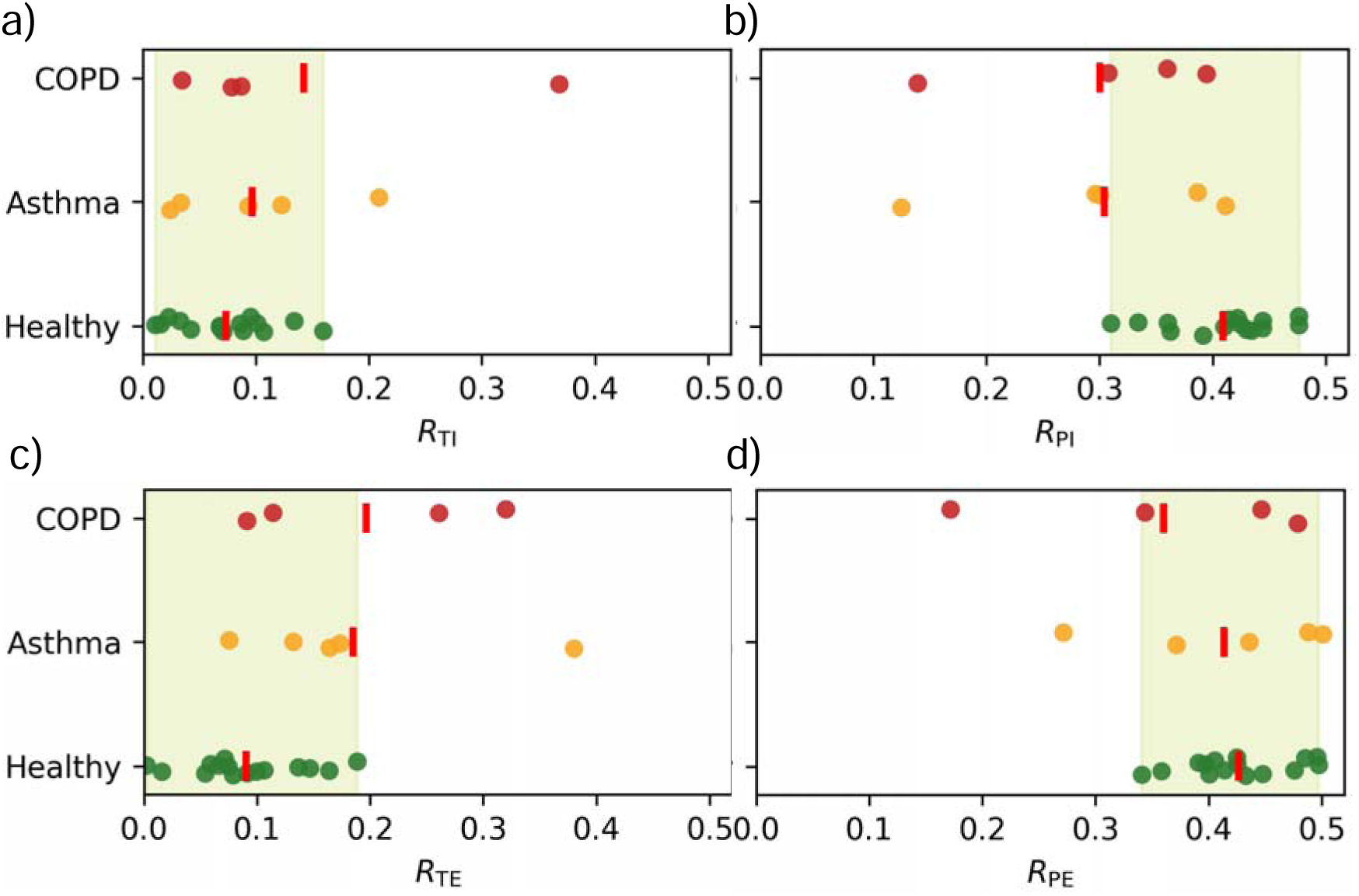
Jittered scatter plots of the phase-resolved integral ratio metrics *R*_TI_ (a), *R*_PI_ (b), *R*_TE_ (c), and *R*_PE_ (d) for healthy subjects and patients with COPD or asthma.

Overall, the proposed phase-resolved integral ratio metrics indicate that healthy subjects exhibit a breathing pattern dominated by quasi-steady phases, whereas patients tend to show a relative shift toward transient-enhanced breathing. This shift is slightly more pronounced as reduced quasi-steady contribution during inhalation and enhanced transient contribution during exhalation.

#### 2.3.2. Conventional Breathing Descriptors

To provide a baseline comparison with the proposed phase-resolved metrics, we analyzed a set of conventional breathing descriptors commonly used in portable or wearable breathing monitoring. These included time-domain breathing metrics (inhalation, exhalation, and breathing cycle durations), amplitude-related metrics (peak, valley, and peak-to-valley amplitudes), and their cycle-to-cycle variability quantified by the coefficient of variation (CV), defined as the ratio of the standard deviation to the mean. In addition, waveform asymmetry was characterized using skewness, and frequency-domain characteristics were evaluated using the relative contribution of high-frequency components.

**Figure 7**a presents jittered scatter plots of breathing cycle duration and peak-to-valley amplitude, together with their corresponding CVs. Other time-domain and amplitude-related metrics obtained from separated inhalation and exhalation phases showed consistent trends and are therefore provided in Figure S3 (Supporting Information). In contrast to the phase-resolved metrics, both breathing duration and amplitude exhibited a broad distribution among healthy participants, indicating substantial inter-individual variability under natural breathing conditions. For the patient groups, breathing cycle duration values were narrowly distributed near the lower bound of the healthy reference range, with one COPD and one asthma subject falling below this range. In comparison, peak-to-valley amplitude values for all patients showed a dispersed distribution but remained fully within the healthy reference range. At the group level, the average amplitude exhibited a gradual decrease from healthy subjects to asthma and further to COPD subjects. Compared with the duration and amplitude, the corresponding CVs displayed a relatively narrow distribution among healthy subjects, defining a stable reference range. Most patients fell within this healthy reference range, with only one COPD subject exceeding the healthy range for the CVs of both duration and amplitude. These results suggest that, on average, patients tend to show faster, shallower, and more variable breathing patterns compared with healthy subjects.

**Figure 7.**
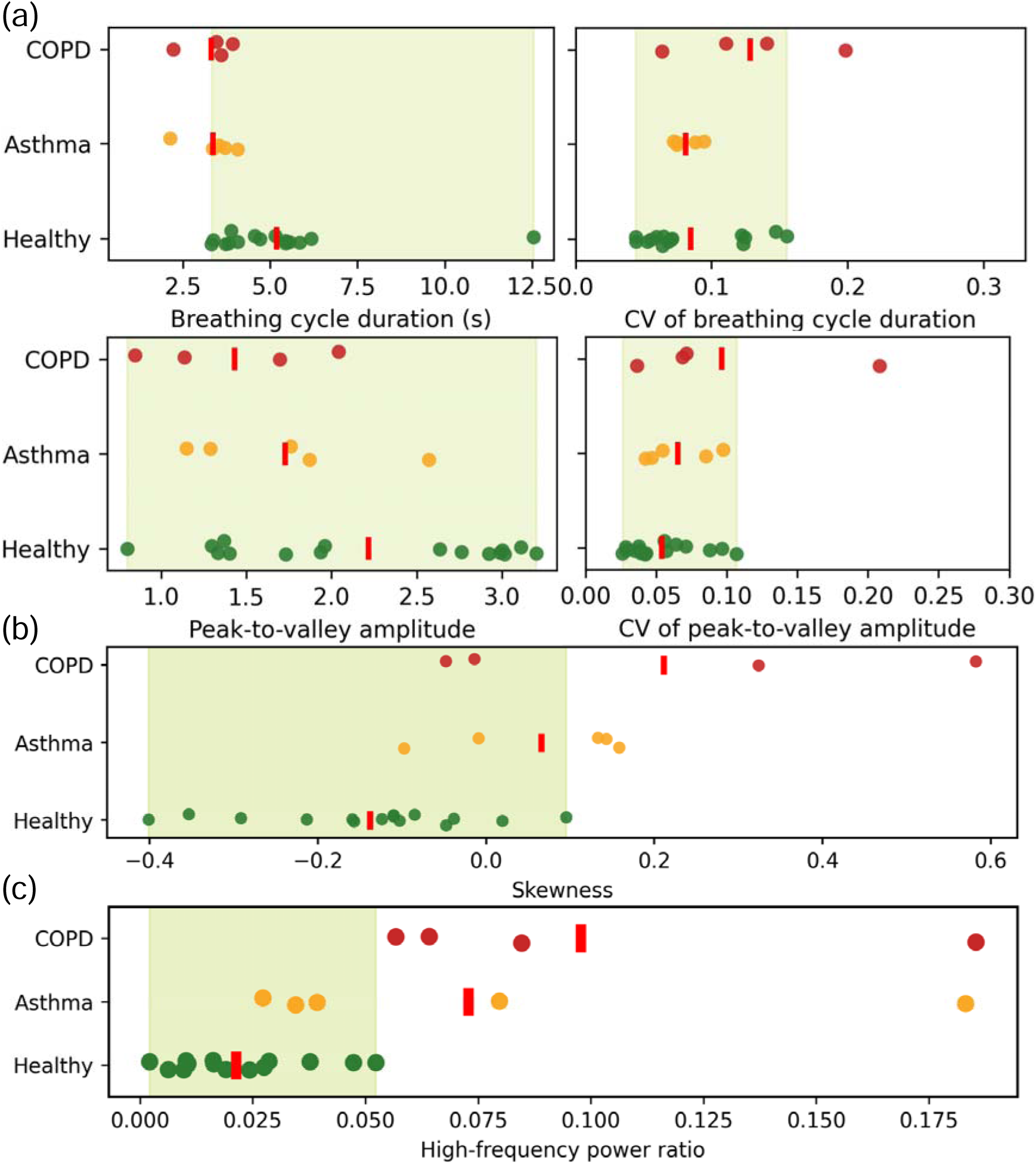
Jittered scatter plots of conventional breathing descriptors for healthy subjects and patients with COPD or asthma. (a) Breathing cycle duration and peak-to-valley amplitude, together with their corresponding cycle-to-cycle coefficients of variation (CV). (b) Skewness of the breathing waveforms. (c) Relative contribution of high-frequency components.

Figures 7b and 7c show jittered scatter plots of skewness and the relative contribution of high-frequency components. FFT amplitude spectra of patients outside the healthy reference range for high-frequency power ratio are provided in Figure S4 (Supporting Information). Skewness values of healthy subjects were distributed over a relatively wide range, with the majority in the negative range, whereas those of patient groups tended to shift toward positive values, with two COPD subjects markedly exceeding both the healthy and asthma ranges. Inspection of the waveforms from the out-of-range subjects revealed that the positive skewness arises from relatively larger quasi-steady exhalation peaks compared with the corresponding inhalation peaks. For the relative contribution of high-frequency components, healthy subjects typically exhibited values below 5%, whereas all COPD subjects and two of the five asthma subjects exceeded this range, with values reaching up to approximately 19%. This indicates the presence of more pronounced rapid intra-cycle fluctuations within individual breathing waveforms in a subset of patients.

#### 2.3.3. Physiological Interpretation and Conceptual Implications

The observed differences across phase-resolved integral metrics and conventional descriptors can be interpreted in the context of known respiratory physiology, which helps clarify their respective information content and inherent limitations.

Under natural breathing conditions, breathing rate, breathing depth, and their cycle-to-cycle variability are primarily influenced by individual breathing habits and momentary physiological conditions. Accordingly, substantial inter-individual variability is observed even among healthy subjects. Although patients showed, at the group level, a tendency toward faster, shallower, and more variable breathing, the considerable overlap with healthy subjects indicates that timing-, amplitude-, and CV-based descriptors alone lack sufficient specificity for reliable discrimination at the individual level. Nevertheless, persistently extreme values, including sustained unusually rapid, shallow, or highly irregular breathing patterns, are more consistent with underlying pathological processes than habitual or behavioral factors.

In contrast, skewness and high-frequency content primarily reflect disturbances in intra-cycle breathing dynamics that arise from the internal structure of the breathing waveform. While shaped by breathing behavior, these descriptors are less amenable to intentional modulation and instead reflect altered inspiratory–expiratory phase distribution and intra-cycle stability. The larger deviations observed in COPD than in asthma are physiologically plausible, given the chronic and largely irreversible nature of airflow limitation in COPD compared with the typically reversible and episodic obstruction in asthma. However, these descriptors summarize overall waveform morphology without explicitly resolving the contributions of individual breathing phases.

The phase-resolved integral metrics further extend this interpretation by explicitly resolving how breathing effort is distributed between transient and quasi-steady phases during inspiration and expiration. The ability to separate these phases is enabled by the characteristic waveform captured by the proposed device, which contains distinct transient and quasi-steady components within each breathing direction. By quantifying the relative contributions of these components, the phase-resolved metrics reveal a shift from quasi-steady–dominated breathing in healthy subjects toward a relatively higher contribution of transient components in patients. Importantly, skewness and high-frequency content can be viewed as indirect manifestations of this same redistribution, whereas the phase-resolved metrics provide a more direct and interpretable representation of it.

Taken together, these results suggest a hierarchy of respiratory information: duration, amplitude, and their variability primarily reflect behavior-driven breathing variability; skewness and high-frequency content capture altered intra-cycle dynamics; and phase-resolved integral metrics directly characterize the underlying redistribution of breathing phases. This conceptual progression highlights the added value of phase-resolved analysis for probing respiratory dynamics that are less accessible through conventional descriptors alone, while remaining physiologically interpretable within a pilot-scale study.

## 3. Conclusion

This work presents a portable breathing monitoring device based on a flexible PZT sensor that simultaneously captures pressure- and temperature-related airflow responses during natural oral breathing. By exploiting polarity-opposed piezoelectric and pyroelectric responses through sensor orientation, the recorded breathing waveforms exhibit a characteristic dual-component structure, comprising a narrow transient spike and a broad quasi-steady peak with opposite polarities during inhalation and exhalation. This intrinsic waveform structure enables direct access to how breathing dynamics are distributed within each breathing cycle, beyond global measures such as breathing rate or depth. On this basis, phase-resolved integral metrics were introduced to quantify the relative contribution of transient and quasi-steady components during inhalation and exhalation. Pilot comparisons among healthy subjects and patients with COPD or asthma show that these metrics capture systematic shifts toward transient-enhanced breathing in patients, providing clearer separation than conventional descriptors based on duration, amplitude, or variability. More broadly, the proposed dual-response sensing approach shifts natural breathing monitoring from behavior-dominated quasi-steady features toward intra-cycle airflow dynamics that are more directly linked to airway mechanics and respiratory system function.

Although the present study is limited in scale and not intended to establish diagnostic criteria, it demonstrates the potential of dual-response, phase-resolved respiratory sensing to access functional changes in natural breathing that are not readily accessible using conventional flow-based monitoring. Future validation in larger and more diverse populations will be essential to establish physiological specificity and to support translation toward practical respiratory monitoring in daily-life settings.

## 4. Materials and Methods

### Sensor Materials and Fabrication

As shown in Figure 1a, the sensor element was encapsulated between two polyimide films for protection. The sensor element consisted of a 20 μm thick fluorophlogopite mica [KMg_3_(AlSi_3_O_10_)F_2_] substrate, a 2 μm thick PbZr_0.52_Ti_0.48_O_3_ (PZT) film, and parallel-plate platinum electrodes (100 nm thick) above and below the PZT layer. The electrodes were fabricated by magnetron sputtering, and the PZT film was formed using a transfer-free sol-gel method.^[42]^ The detailed fabrication process followed those reported in our previous work.^[38]^

### Signal Conditioning and Data Acquisition

The electrical charges generated by the sensor were converted into voltage signals using an onboard charge amplifier with a 1.65 V bias applied, resulting in an output voltage range of 0–3.3 V. In subsequent signal processing, this bias offset was subtracted, and the voltage waveforms were treated as effective bipolar signals in the range of ±1.65 V. The amplified signals were digitized by an onboard 12-bit analog-to-digital converter and transmitted wirelessly via Bluetooth to an iOS-based mobile terminal for recording. The sampling interval was 3.2 ms. All circuit configurations and acquisition parameters followed those reported in our previous work.^[38]^

### Setup and Protocols of Breathing Measurements

Breathing measurements were performed in both healthy volunteers and patients with diagnosed respiratory diseases using a unified experimental protocol. To accommodate the slightly larger outer diameter (26 mm) of the mouthpieces used in the hospital, the inner diameter of the flow channel in the proposed PZT-based device used for patient measurements was slightly increased compared with that used for healthy subjects, while all other components and acquisition settings were identical. The characteristic waveform features were consistently observed across both device form factors. For measurements in healthy volunteers conducted in a laboratory setting, a commercially available oral respiratory flow sensor (SFM3300-AW, Sensirion, Figure 3a) was used to simultaneously record airflow rate signals (100 Hz) on a personal computer. Due to practical constraints in the clinical environment, simultaneous flow sensor measurements were not performed during patient measurements.

All measurements, except for specifically noted control experiments, were conducted under seated, resting conditions in an indoor environment at room temperature. Participants were instructed to breathe naturally through a mouthpiece connected to the proposed device. To minimize voluntary modulation of breathing, visual feedback of the recorded signals was not provided during the measurements. Each recording lasted approximately 60 s, which was sufficient to capture multiple consecutive breathing cycles under stable conditions. For healthy volunteers, the use of a nasal clip was determined on an individual basis depending on their ability to maintain natural oral breathing; for patient measurements, a nasal clip was consistently used.

All breathing measurements were conducted in accordance with the Declaration of Helsinki and were approved by the Graduate School of Informatics, Nagoya University (approval number: hc24-12), as well as by the institutional review board of the participating hospital. Written informed consent was obtained from all participants prior to participation. *Phase Segmentation of Breathing Cycles*: Each breathing cycle was segmented into four phase-resolved regions used to compute *R*_TI_, *R*_PI_, *R*_TE_, and *R*_PE_. Segmentation was based on characteristic features of the voltage waveform. The onset of inhalation or exhalation was first identified, appearing as a local minimum or maximum, respectively, reflecting signal polarity reversal at the phase transition. When clear local extrema were absent, the onset was identified as an inflection point characterized by a pronounced change in signal slope accompanying polarity reversal. If multiple inflection points were present, the one immediately preceding the spike feature was selected, as the spike consistently occurred shortly after the true phase onset. Owing to subject-dependent waveform variations and occasional local noise, inhalation and exhalation onsets were determined by visual inspection to ensure consistent and physiologically reasonable phase assignment across recordings. The transient phase was defined as the interval from the inhalation or exhalation onset to the subsequent zero-crossing of the voltage signal (0 V). The remaining interval until the onset of the next inhalation or exhalation phase was defined as the quasi-steady phase.

### Frequency-Domain Analysis

The relative contribution of high-frequency components was calculated by Fourier analysis of the breathing waveforms. Prior to transformation, a Hann window was applied to reduce spectral leakage. The frequency range was set to 0.05–2.0 Hz, divided into a low-frequency band (0.05–0.6 Hz) and a high-frequency band (0.6–2.0 Hz). The boundary frequency of 0.6 Hz was chosen to remain above the fundamental breathing frequency even under fast breathing conditions, while avoiding contamination from higher-order harmonics.

## Supporting Information

Supporting Information, including notes and figures, is available on medRxiv. Movie S1 is not publicly available due to ethical and privacy considerations and can be provided by the corresponding author upon reasonable request.

## Acknowledgements

This work was supported in part by the Grant-in-Aid for JSPS Fellows (No. 24KJ1281), the FY2023 University-Based New Industry Creation Fund Project Startup Ecosystem Co-Creation Program Gap Fund Program “Step 1”, and JSPS KAKENHI Grant Number 25K22887.

## Conflict of Interest

The authors declare no conflict of interest.

## Data Availability Statement

The data that support the findings of this study are available from the corresponding author upon reasonable request.

## Notes

### Competing Interest Statement

The authors have declared no competing interest.

### Author Declarations

The Ethics Committee of the Graduate School of Informatics and the relevant ethics review bodies at the Higashiyama Campus, Nagoya University gave ethical approval for this work. The Ethics Committee of Daido Hospital also approved this work.

