## Supporting Information for "Portable Breathing Monitoring with Phase-Resolved Airflow Dynamics Enabled by a Dual-Response Flexible PZT Sensor"

T. Uchiyama
Institute of Materials and Systems for Sustainability, Nagoya University
Furo-cho, Chikusa-ku, Nagoya 464-8601, Japan

K. Yoshikawa
Department of Respiratory Medicine, Daido Hospital, Kojunkai Social Medical Corporation
9 Hakusui-cho, Minami-ku, Nagoya 457-8511, Japan

T. Mano
Chuo Graduate School of Strategic Management, Chuo University742-1 Higashinakano, Hachioji-shi, Tokyo 192-0393 Japan

### Note S1. Comparison of Pyroelectric Sensitivity between Flexible PZT and PVDF Sensors

The pyroelectric sensitivity of the flexible PZT was compared with that of a commercially available polyvinylidene difluoride (PVDF) sensor. A cyclic temperature variation of the sensor surface between 25.0 and 36.4 °C over 2 s was applied using the same thermal excitation method as illustrated in Figure 2b of the main text. Except for replacing the sensor, all other conditions were kept identical. The average peak-to-valley pyroelectric amplitude was 0.37 ± 0.039 V for the PZT sensor and 0.59 ± 0.081 V for the PVDF sensor, indicating comparable pyroelectric sensitivity (**Figure S1**).

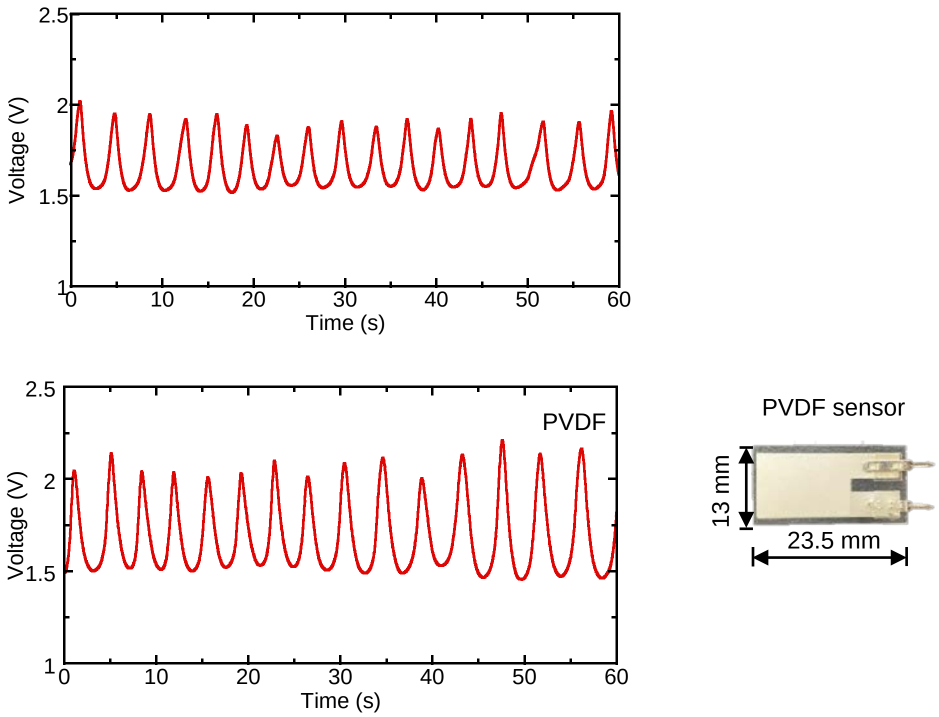


**Figure S1.** Pyroelectric voltage responses of flexible PZT (a) and PVDF (b) sensors measured under identical thermal excitation. A photograph of the PVDF sensor is shown in (b).

**Note S2. Neutral Plane Calculation of the Flexible PZT Sensor**

The position of the neutral mechanical plane of the flexible PZT sensor was calculated based on its multilayer structure (Figure 1a in the main text and **Table S1**) using the following equation:

$$y_{\mathrm{N}} = \frac{\sum_{i=1}^{n} E_i b_i t_i y_i}{\sum_{i=1}^{n} E_i b_i t_i} \tag{S1}$$

where $E_i$, $b_i$, $t_i$ are the Young's modulus, width, and thickness of each layer, respectively, and $y_i$ is the distance from the center plane of each layer to the top surface of the first layer. Using the values list in Table S1, the neutral plane was calculated to be approximately 15.1 μm from the top surface of the first layer, lying within the mica layer.

**Table S1** Material, thickness, width, and Young's modulus of each layer of the flexible PZT sensor. The Young's modulus values were taken from Refs. [1–4].

| Layer No. | Material | Thickness (μm) | Width (mm) | Young's modulus (GPa) |
|---|---|---|---|---|
| 1 | Polyimide film | 10.0 | 15.0 | 2.5 |
| 2 | Pt | 0.1 | 10.0 | 116 |
| 3 | PZT | 2.0 | 10.0 | 140 |
| 4 | Pt | 0.1 | 10.0 | 116 |
| 5 | Mica | 20.0 | 10.0 | 5.4 |
| 6 | Polyimide film | 10.0 | 15.0 | 2.5 |

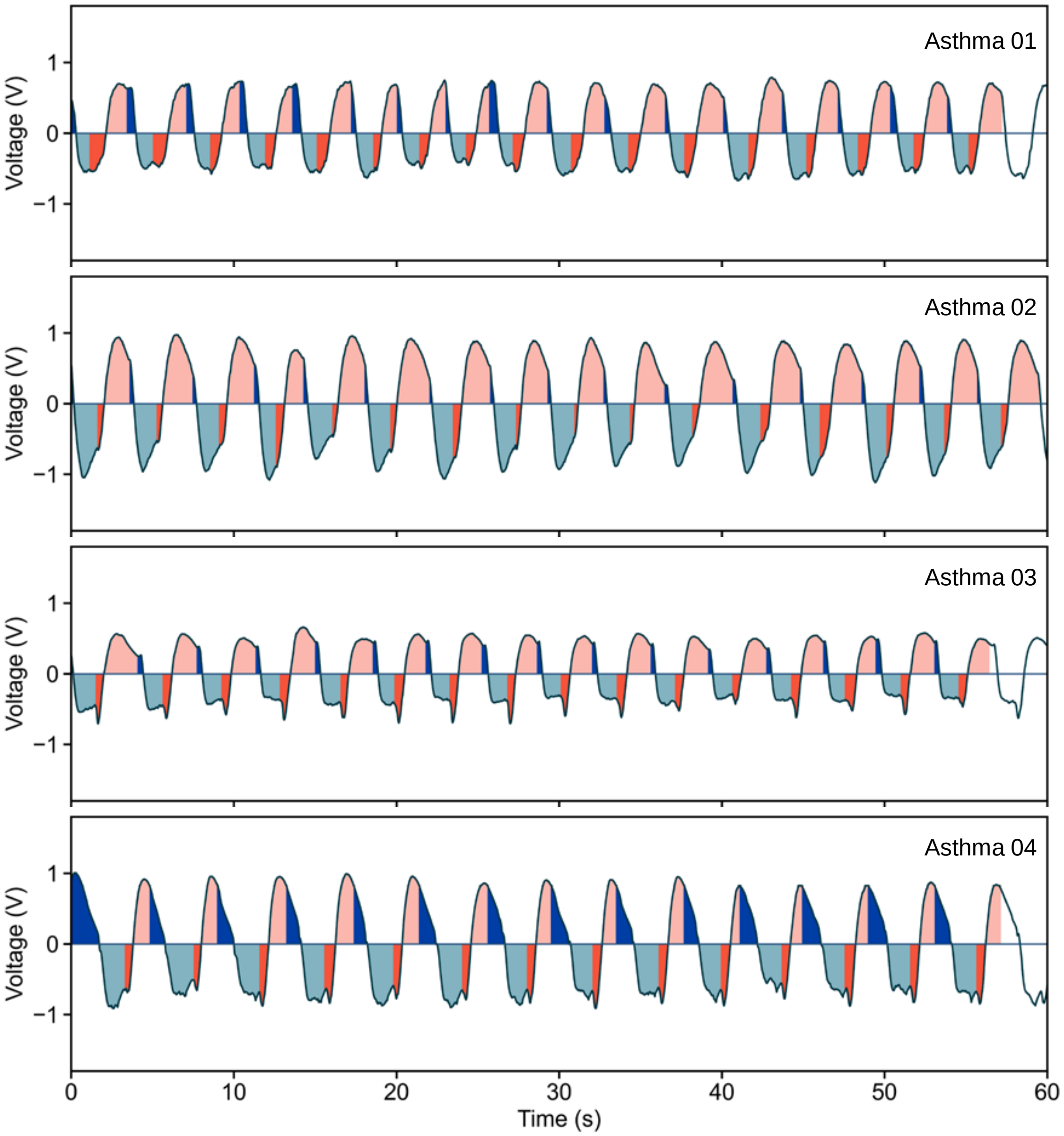

Asthma 01
Asthma 02
Asthma 03
Asthma 04
Voltage (V)
1
0
−1
0
10
20
30
40
50
60
Time (s)

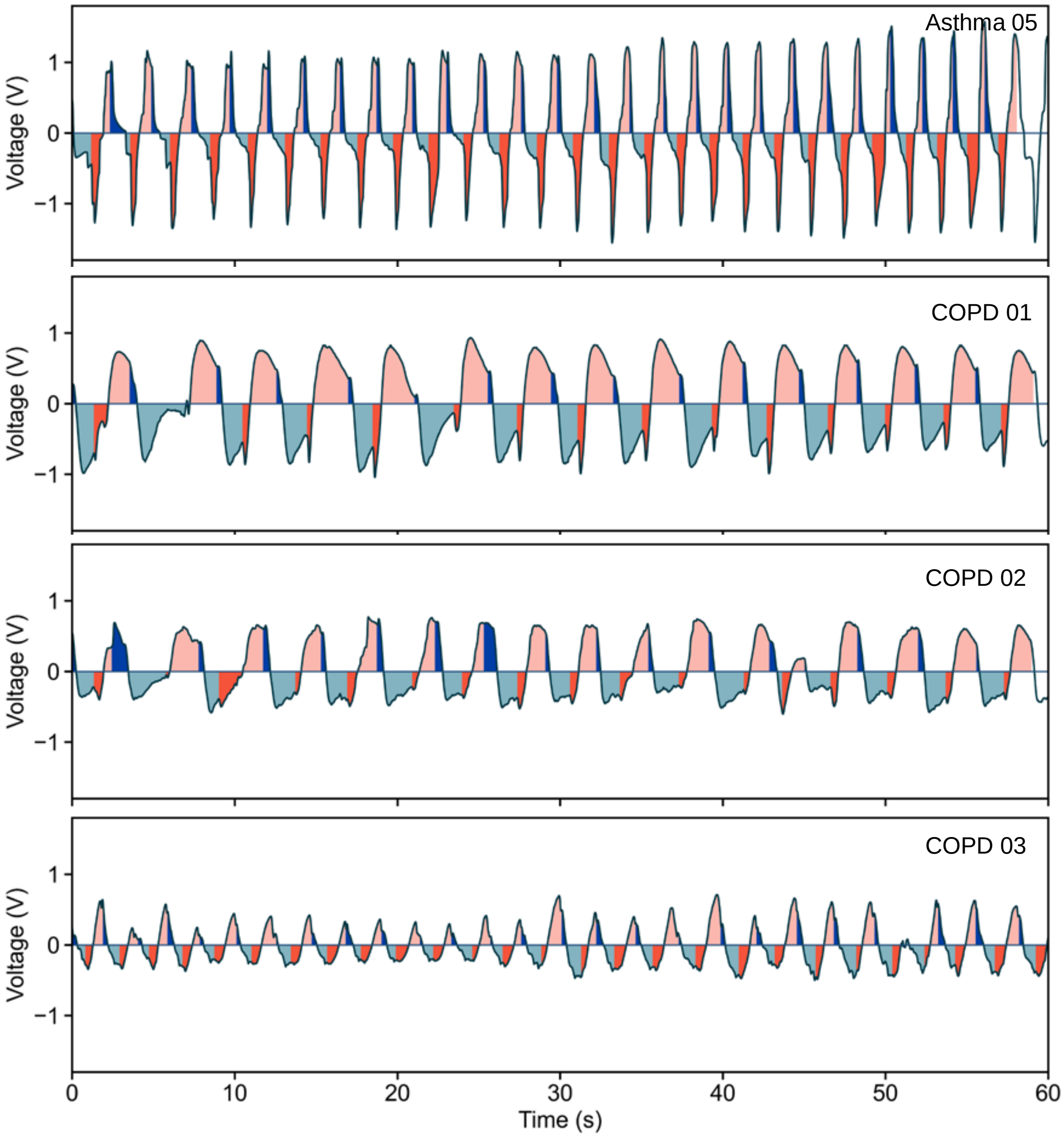


**Figure S2.** Breathing waveforms from eight patients measured by the proposed device. The waveform of the remaining patient is shown in Figure 5 of the main text.

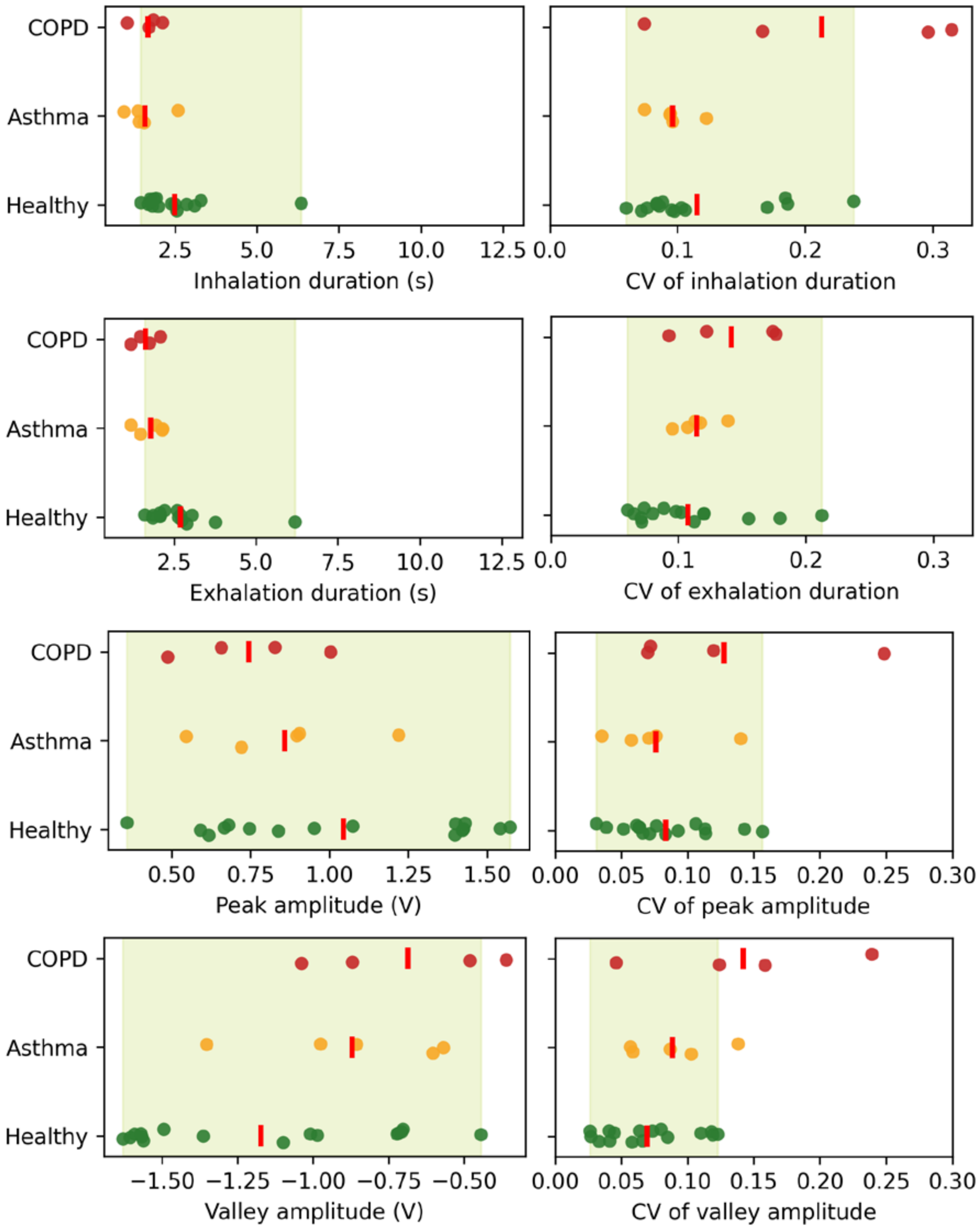


**Figure S3.** Jittered scatter plots of inhalation duration, exhalation duration, peak amplitude, valley amplitude, together with their corresponding cycle-to-cycle coefficients of variation (CV), for healthy subjects and patients with COPD or asthma.

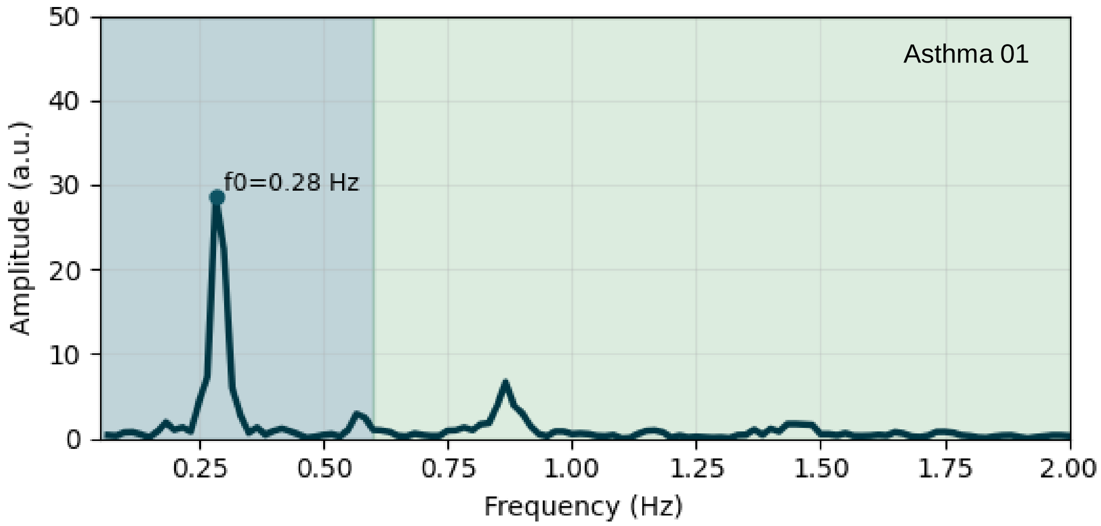
Asthma 01
f0=0.28 Hz
Amplitude (a.u.)
Frequency (Hz)
0
10
20
30
40
50
0.25
0.50
0.75
1.00
1.25
1.50
1.75
2.00

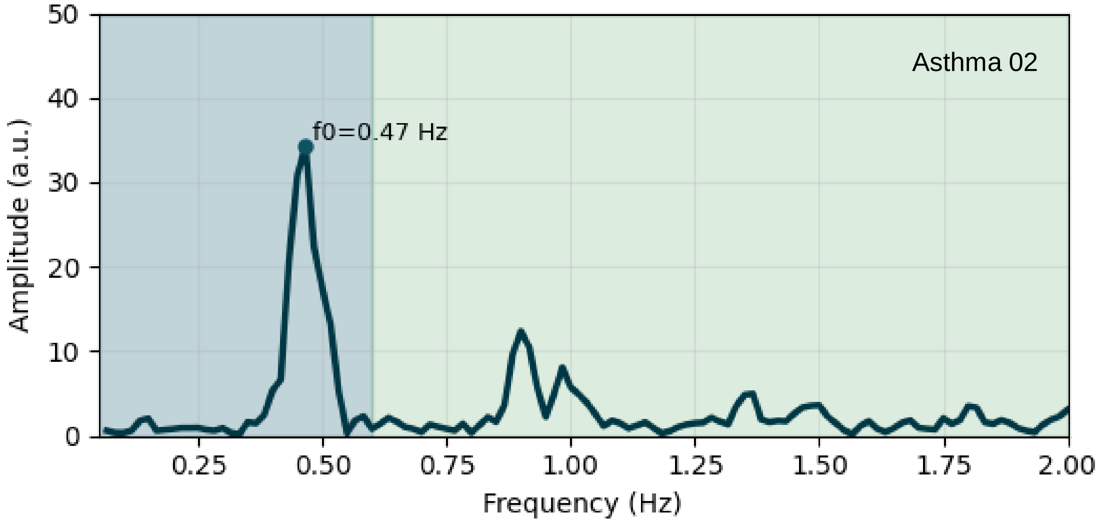
Asthma 02
f0=0.47 Hz
Amplitude (a.u.)
Frequency (Hz)
0
10
20
30
40
50
0.25
0.50
0.75
1.00
1.25
1.50
1.75
2.00

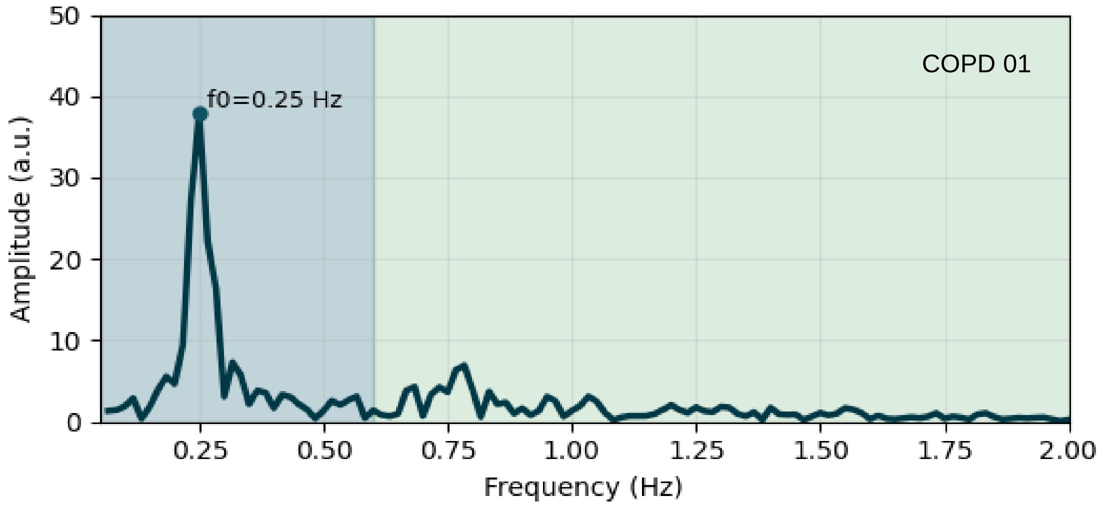
COPD 01
f0=0.25 Hz
Amplitude (a.u.)
Frequency (Hz)
0
10
20
30
40
50
0.25
0.50
0.75
1.00
1.25
1.50
1.75
2.00

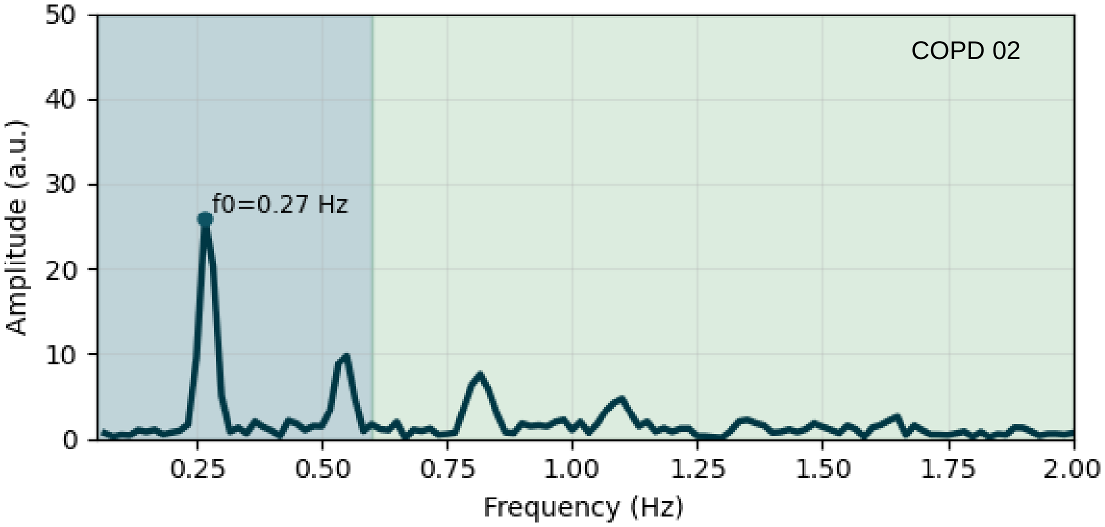
COPD 02
f0=0.27 Hz
Amplitude (a.u.)
Frequency (Hz)
0
10
20
30
40
50
0.25
0.50
0.75
1.00
1.25
1.50
1.75
2.00

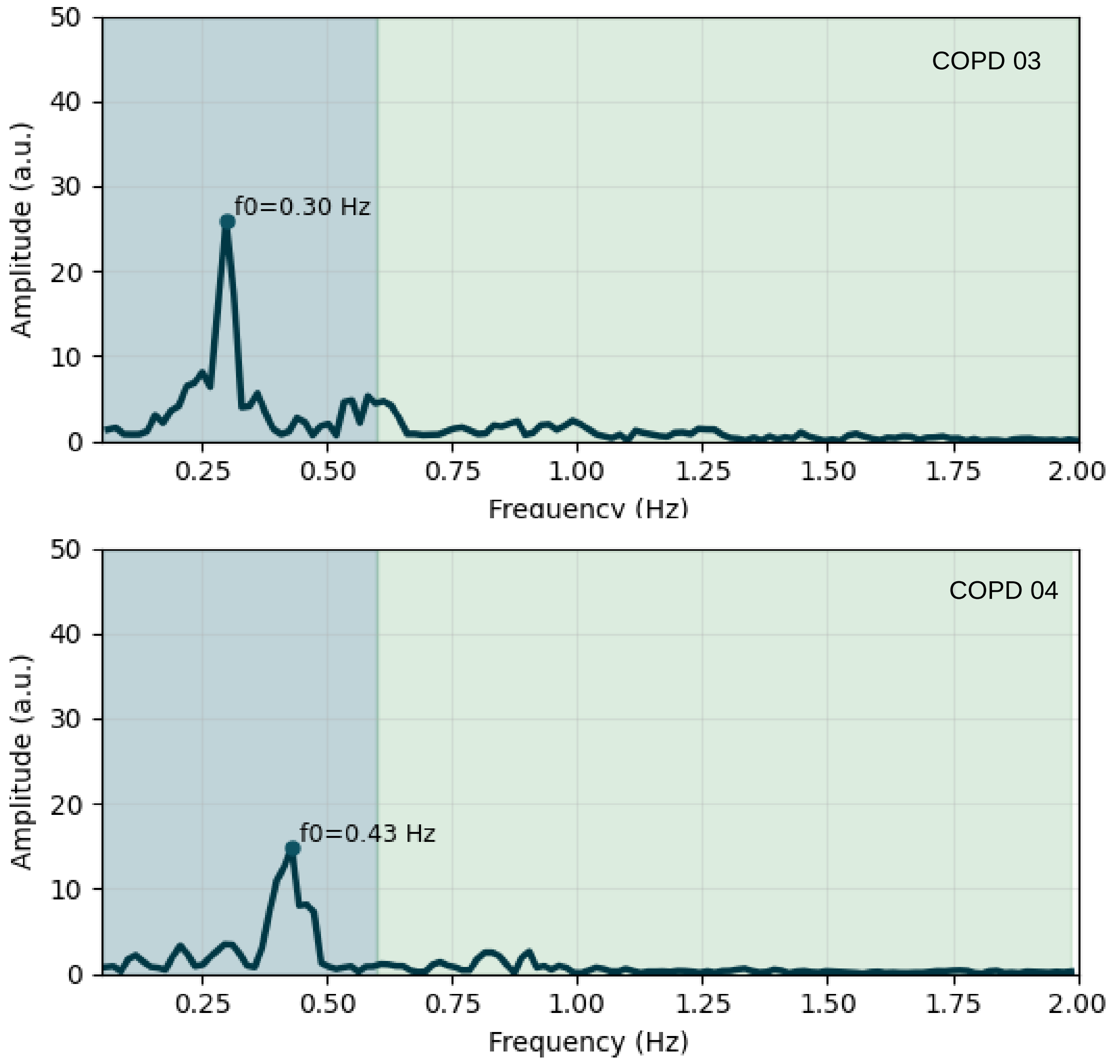


**Figure S4.** FFT amplitude spectra of patients whose high-frequency power ratios fall outside the healthy reference range, as defined in Figure 7c of the main text.
